# COVID-19: impact of original, Gamma, Delta, and Omicron variants of SARS-CoV-2 in vaccinated and unvaccinated pregnant and postpartum women

**DOI:** 10.1101/2022.10.05.22280754

**Authors:** Fabiano Elisei Serra, Elias Ribeiro Rosa, Patricia de Rossi, Rossana Pulcineli Vieira Francisco, Agatha Sacramento Rodrigues

## Abstract

**Introduction:** This study compares the clinical characteristics and disease progression of vaccinated and unvaccinated pregnant and postpartum women positive for the original, Gamma, Delta, and Omicron variants of severe acute respiratory syndrome coronavirus 2 (SARS-CoV-2) using Brazilian epidemiological data.

**Methods:** Data of pregnant or postpartum patients with coronavirus disease 2019 (COVID-19) SARS-CoV-2 confirmed using polymerase chain reaction from February 2020 to July 2022 were extracted from a Brazilian national database. The patients were divided based on vaccination status and viral variant (original, Gamma, Delta, and Omicron). The patients’ demographic data, clinical characteristics, comorbidities, signs, symptoms, and outcomes were retrospectively compared.

**Results:** Data from 10,003 pregnant and 2,361 postpartum women were extracted from the database. Among unvaccinated patients, postpartum women were more likely to be admitted to the intensive care unit (ICU). These patients were more likely to require invasive ventilation when infected with the original, Gamma, and Omicron variants and were more likely to die when infected with the original and Gamma variants. Patients who were vaccinated had reduced adverse outcomes including ICU admission, requirement for invasive ventilation, and death.

**Conclusion:** Postpartum women were more likely to develop severe COVID-19 that required ICU admission or invasive ventilatory support or led to death, among all variants, especially when the patients were unvaccinated. Therefore, the risk of severe COVID-19 should not be underestimated after delivery. Vaccinated patients had a lower risk of severe outcomes. Vaccination should be a top priority in pregnant and postpartum patients.

**WHAT IS ALREADY KNOWN ON THIS TOPIC:** The obstetric population has a higher risk of adverse outcomes due to coronavirus disease 2019 (COVID-19). Few studies have compared the outcomes of pregnant and postpartum patients or vaccinated and unvaccinated patients; however, no studies have separately investigated the effects of each viral variant.

**WHAT THIS STUDY ADDS:** Postpartum women are more likely to have adverse outcomes, including the requirements for intensive care and invasive ventilation and death, compared with pregnant women. Vaccinated women had fewer adverse outcomes. The viral variants did not significantly affect the outcomes of these patients.

**HOW THIS STUDY MIGHT AFFECT RESEARCH, PRACTICE, OR POLICY:** The risks of COVID-19 infection should not be underestimated in postpartum women. Postpartum women infected with COVID-19, especially those who are not vaccinated, should be monitored carefully. Vaccination should be a top priority in pregnant and postpartum women.

## INTRODUCTION

Coronavirus disease 2019 (COVID-19) is caused by severe acute respiratory syndrome coronavirus 2 (SARS-CoV-2), which was first reported in Wuhan, China in December 2019. COVID-19 was declared a pandemic in March 2020.[1] As of 26 August 2022, over 596 million cases and 6.4 million deaths have been reported worldwide.[2]

COVID-19 affects several systems, with a clinical spectrum ranging from asymptomatic to severe and critical disease. Morbidity and mortality among pregnant women and perinatal outcomes are significant concerns. Initial studies did not report a higher mortality rate or more severe disease in obstetric patients with COVID-19; however, later studies reported that obstetric patients had a higher risk of intensive care admission, requiring invasive ventilation, and death, compared with nonpregnant women.[3-11]

During the course of the pandemic, SARS-CoV-2 developed mutations, resulting in new variants. Variants of concern (VOCs) have higher transmissibility, increased virulence, and altered COVID-19 epidemiology and presentation. The effectiveness of public health, social measures, and available therapeutics decreases for VOCs.[12] As of July 30, 2022, three VOCs have been detected in Brazil: the Gamma, Delta, and Omicron variants.[13]

A high number of maternal deaths due to COVID-19 has been reported in Brazil, including 124 deaths as of 18 June 2020,[14] and 295 maternal deaths have been confirmed using reverse transcriptase polymerase chain reaction (RT-PCR) as of 2 January 2021. Puerperal women have a higher risk of serious outcomes (such as intensive care unit (ICU) admission, requirement for invasive and non-invasive ventilatory support, and death) than pregnant women do.[15] The Brazilian Obstetric Observatory for COVID-19 was created on 7 April 2021, and it provides updated data released by the Ministry of Health of Brazil. In total, 1,031 maternal deaths due to COVID-19 (including 680 pregnant women and 351 postpartum women) were reported in Brazil as of 5 May 2021.[16] Even after vaccination campaigns began for pregnant and postpartum women in Brazil on 1 May 2021, pregnant and postpartum women continued to have higher risks of admission to the ICU, intubation, and death, compared with non-pregnant women and men.[17, 18]

As vaccination is one of the most effective strategies to reduce the transmission and severity of infectious diseases, it is important to evaluate not only the effectiveness of the vaccines in the obstetric population but also their protection against the new VOCs.[19, 20] Therefore, this study compares the clinical characteristics and outcomes of pregnant and postpartum women based on vaccination status and VOC, using data from a Brazilian national database.

## METHODS

Patient data were extracted from the System of Information about Epidemiological Surveillance of Influenza (SIVEP-Gripe), a Brazilian national database that includes surveillance data regarding severe acute respiratory syndrome (SARS).[21] Compulsory reporting of SARS due to the flu (acute respiratory conditions, characterized by at least two of the following signs and symptoms: fever, chills, headache, sore throat, cough, coryza, anosmia, and ageusia), associated with dyspnoea, persistent chest pressure, oxygen saturation (SpO_2_) < 95% on room air, or cyanosis is included in this database. All public and private hospitals must report all hospitalizations and deaths due to SARS-CoV-2.

The SIVEP-Gripe includes demographic data (sex, age, skin colour/ethnicity, level of education, obstetric status, and city of residence), clinical data (signs and symptoms, risk factors, and comorbidities), epidemiological data (previous flu vaccinations, community-acquired infection, or nosocomial infection), and laboratory and etiological diagnoses. Hospital and ICU admission, requirement for ventilatory support (invasive or non-invasive), and disease outcome (death or cure) are also reported. SIVEP-Gripe records are publicly-available, anonymized data. Therefore, according to Brazilian Ethics regulatory requirements, ethical approval by an Institutional Review Board was not necessary for this study. This study followed the principles of the Declaration of Helsinki.

In this study, the database was searched from 29 December 2019 to 16 July 2022 (epidemiological weeks 1-53 in 2020; 1-52 in 2021; and 1-28 in 2022), as the first Brazilian records began in epidemiological week 8 (symptom onset for the first confirmed case of COVID-19 in Brazil occurred on 17 February 2020).[22] The search included all records of obstetric patients aged 10 to 49 years, who were hospitalized with COVID-19 (confirmed using RT-PCR for SARS-CoV-2) (Figure 1). A total of 12,364 women were divided into two groups: pregnant (n=10,003) and puerperal (n=2,361). Each group was subdivided based on the dominant VOCs in Brazil at the time of diagnosis: original, Gamma, Delta, and Omicron. The Gamma variant was considered dominant from 1 February 2021 to 31 July 2021, the Delta variant from 1 August 2021 to 31 December 2021, and the Omicron variant from 1 January 2022 to 16 July 2022.[13] Vaccination of pregnant and postpartum patients began in May 2021 in Brazil, during the dominance of the Gamma variant.[18] Therefore, patients in the Gamma, Delta, and Omicron subgroups were also divided based on vaccination status. Only valid responses of the analysed variable were considered. The tables of analyses show the number of valid observations for each variable. Variables used in the analysis were age, skin colour/ethnicity, level of education, comorbidities, signs and symptoms, admission to the ICU, requirement for invasive respiratory support, and outcome (death). The reported comorbidities included chronic cardiovascular disease, asthma, obesity, and diabetes. The reported signs and symptoms included fever, cough, sore throat, dyspnoea, respiratory discomfort, SpO_2_ < 95% on room air, diarrhoea, vomiting, abdominal pain, fatigue, anosmia, and ageusia.

**Figure 1.**
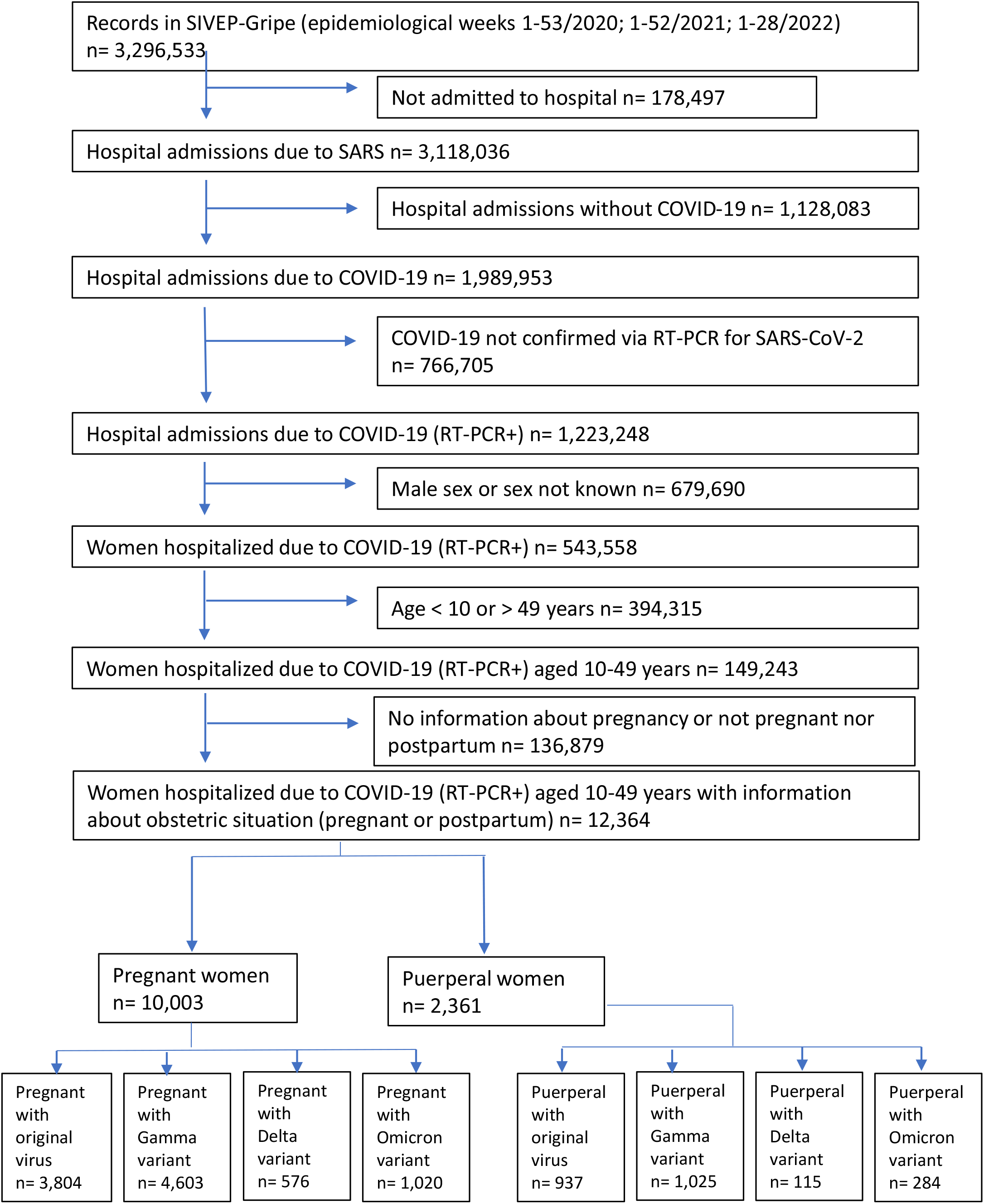
Study Profile: case selection flowchart. **Abbreviations:** SIVEP-Gripe: System of Information about Epidemiological Surveillance of Influenza; SARS: severe acute respiratory syndrome; COVID-19: Coronavirus disease 2019; RT-PCR: polymerase chain reaction

### data analysis

Categorical variables are presented as numbers (n) and percentages (%). These were compared using the chi-square or Fisher’s exact test. The odds ratio (OR) and 95% confidence interval (CI) were calculated to compare the relative odds of the occurrence of a variable of interest between the groups. All statistical analyses were conducted using R software (version 4.0.3; R Foundation for Statistical Computing Platform).[23]

## RESULTS

Data of 12,364 obstetric women (age range: 10-49 years), who were hospitalized with COVID-19 confirmed using RT-PCR for SARS-CoV-2, were included in this study. The pregnant group included 10,003 patients, while the puerperal group included 2,361 patients. Patient age and education level were not significantly different between the groups (Table 1). The rates of the Omicron variant in different ethnic groups were significantly different between the pregnant and puerperal groups. Patients in the puerperal group had fewer comorbidities.

**Table 1.**
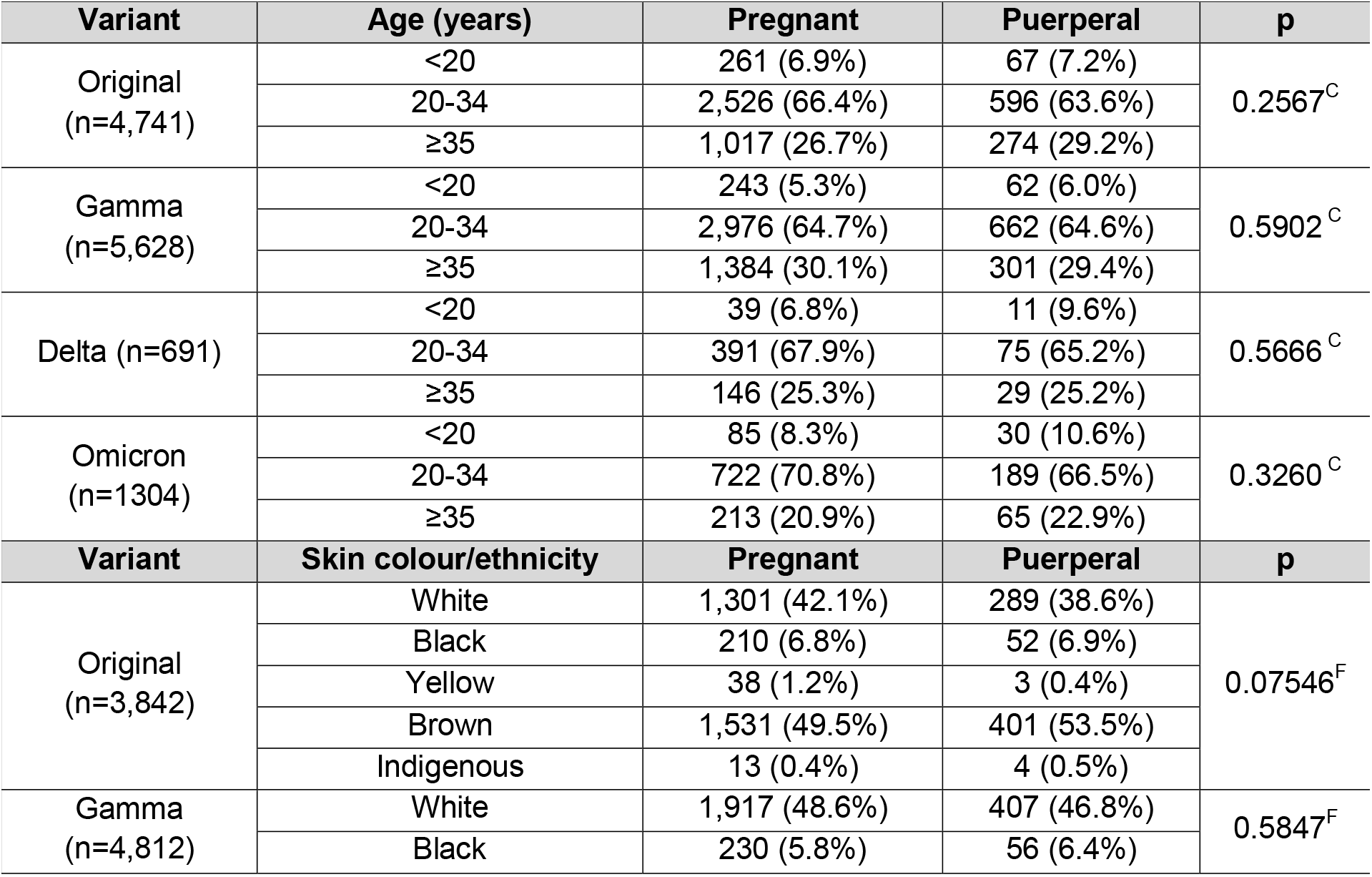
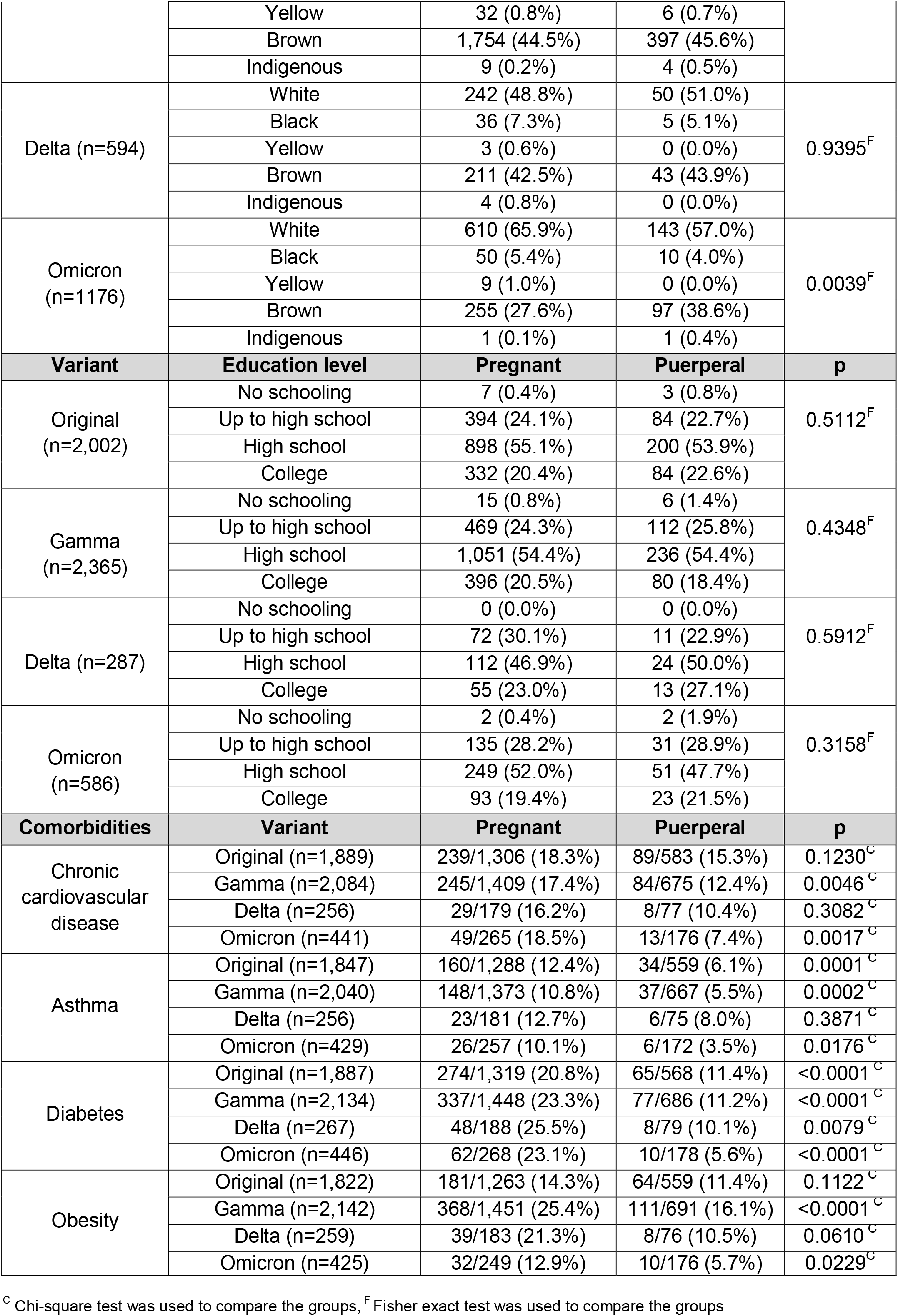
Patient characteristics.

Among unvaccinated patients who were positive for the original SARS-CoV-2 variant, the puerperal group, compared with the pregnant group, had a lower frequency of fever (OR: 0.76; 95% CI: 0.65 – 0.90), coughing (OR: 0.69; 95% CI: 0.58 – 0.83), vomiting (OR: 0.47; 95% CI: 0.33 – 0.65), and ageusia (OR: 0.72; 95% CI: 0.53 – 0.97) and a higher frequency of respiratory discomfort (OR: 1.19; 95% CI: 1.01 – 1.39), SpO_2_ < 95% on room air (OR: 1.70, 95% CI: 1.44 – 2.00), ICU admission (OR: 1.82; 95% CI 1.54 – 2.13), invasive respiratory support (OR: 2.62; 95% CI 2.13 – 3.22), and death (OR: 2.48; 95% CI: 1.96 – 3.13) (Tables 2 and 3). Unvaccinated patients in the puerperal group who had the Gamma variant had a lower frequency of fever (OR: 0.61; 95% CI: 0.50 – 0.76), coughing (OR: 0.56; 95% CI: 0.44 – 0.71), dyspnoea (OR: 0.75; 95% CI: 0.60 – 0.94), vomiting (OR: 0.49; 95% CI: 0.32 – 0.73), anosmia (OR: 0.60; 95% IC 0.43 – 0.82), and ageusia (OR: 0.69; 95% CI: 0.50 – 0.94); however, they were more likely to be admitted to the ICU (OR: 1.58; 95% CI: 1.29 – 1.93), require invasive respiratory support (OR: 2.17; 95% CI: 1.75 – 2.70), and die (OR: 1.99; 95% CI: 1.56 – 2.53), compared with unvaccinated pregnant patients. There were no differences in symptoms, ICU admission, requirement for invasive respiratory support, or death between the two groups among unvaccinated patients with the Delta variant. Unvaccinated patients in the puerperal group with the Omicron variant had a lower frequency of vomiting (OR: 0.23; 95% CI: 0.03 – 0.98) and a higher frequency of ICU admission (OR: 2.61; 95% CI: 1.25 – 5.38) and invasive respiratory support (OR: 3.09; 95% CI: 1.12 – 8.50), compared with unvaccinated pregnant patients with the Omicron variant.

**Table 2.**
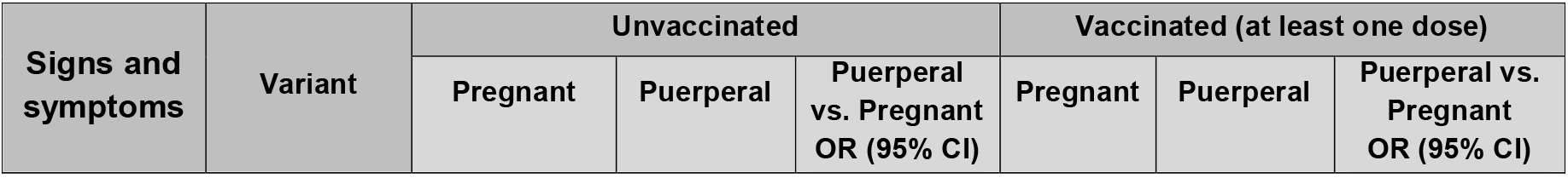
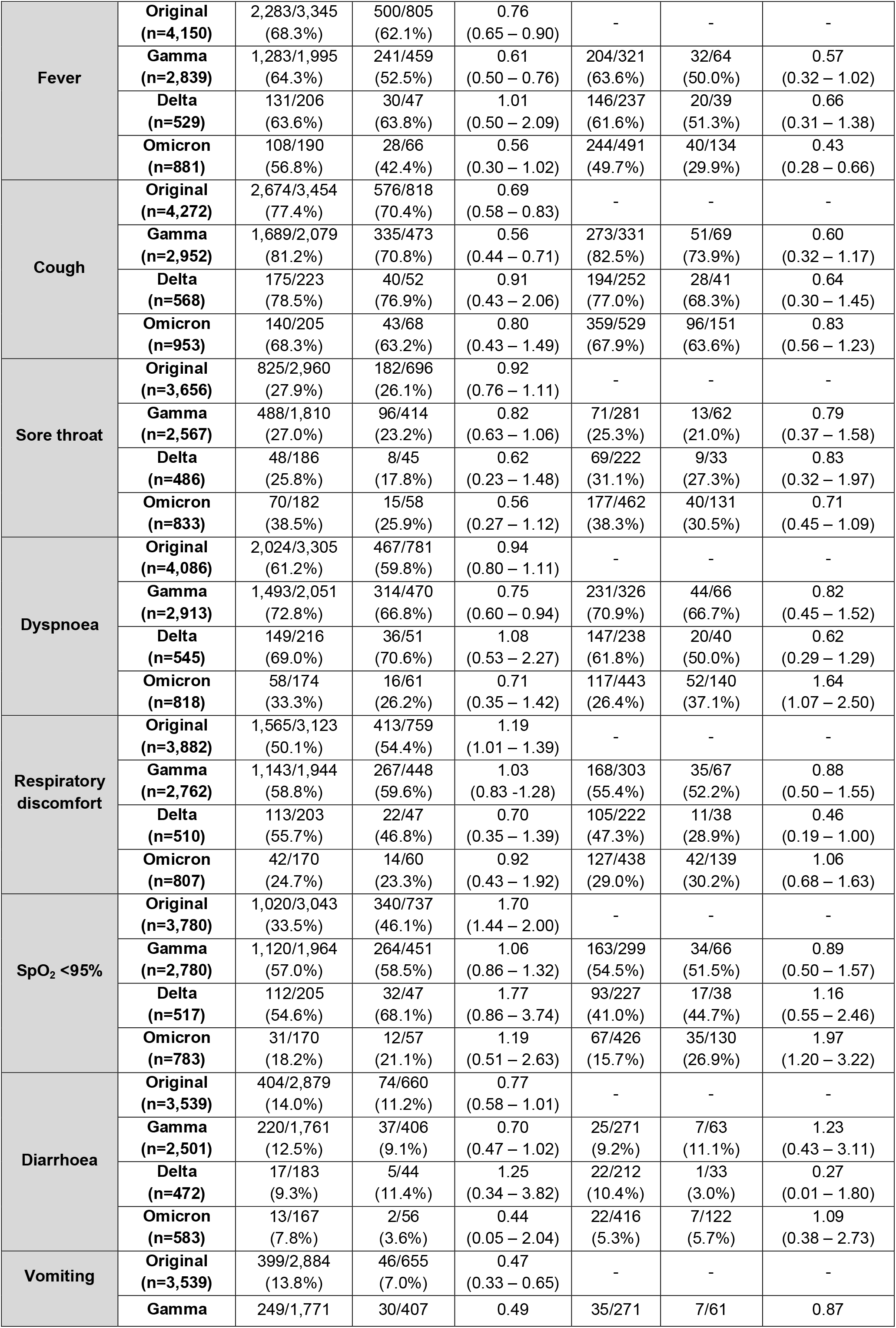
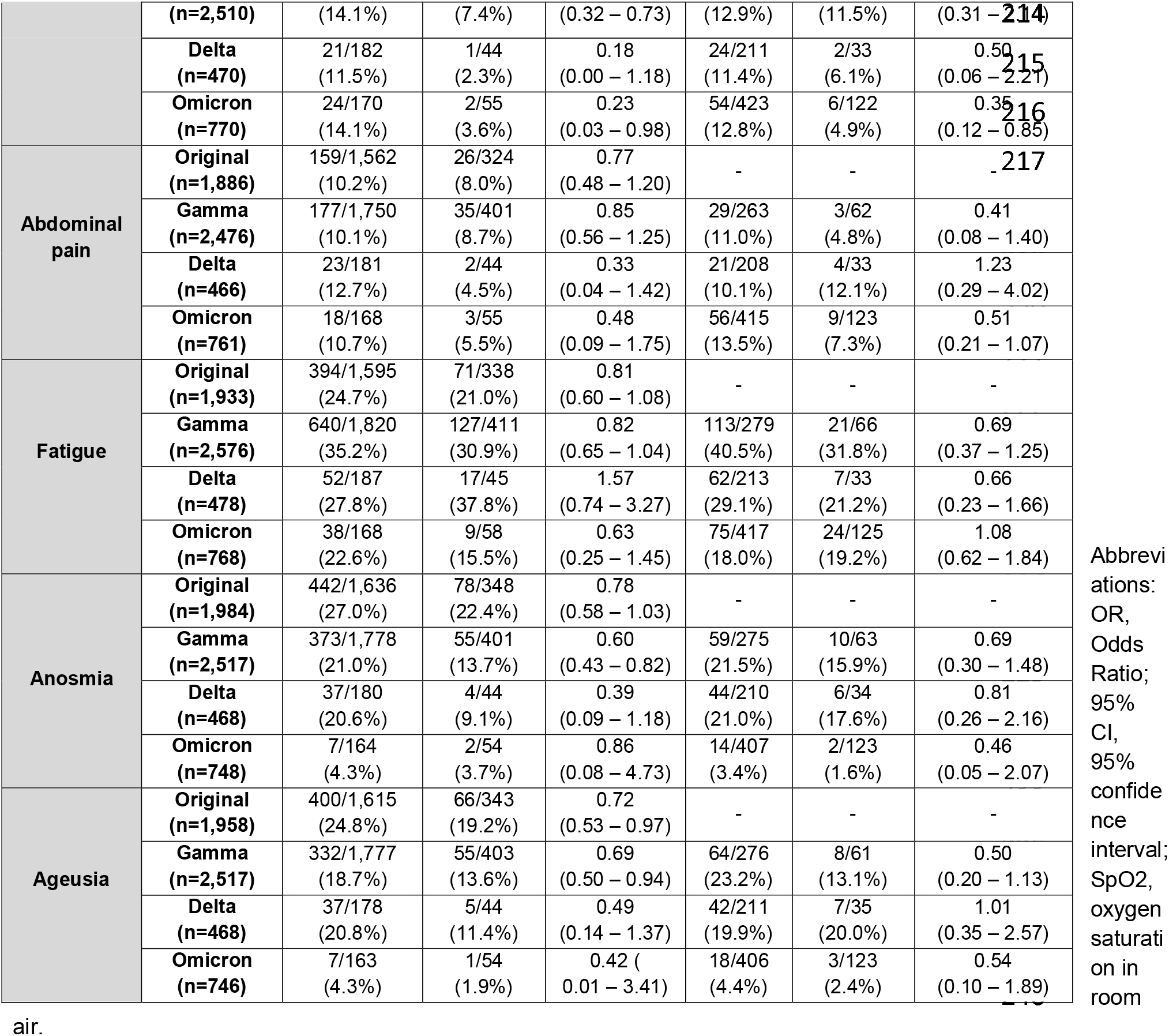
Signs and symptoms of COVID-19 in pregnant and postpartum women.

**Table 3.**
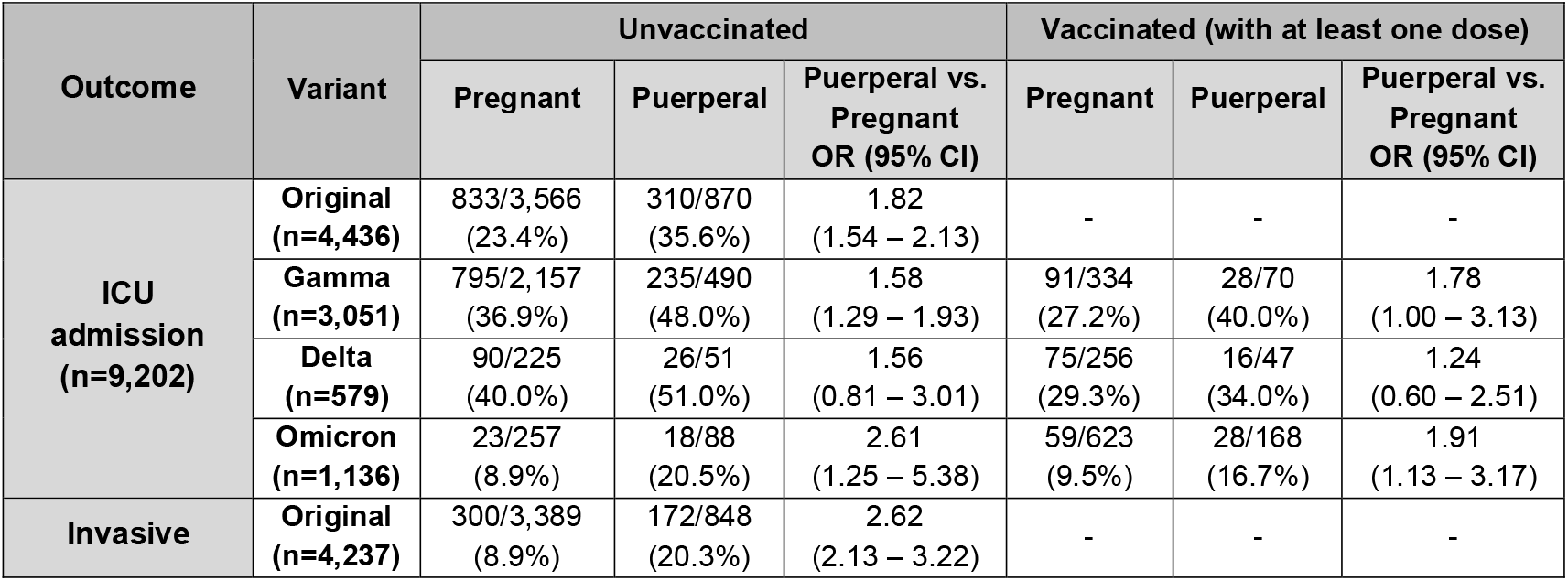
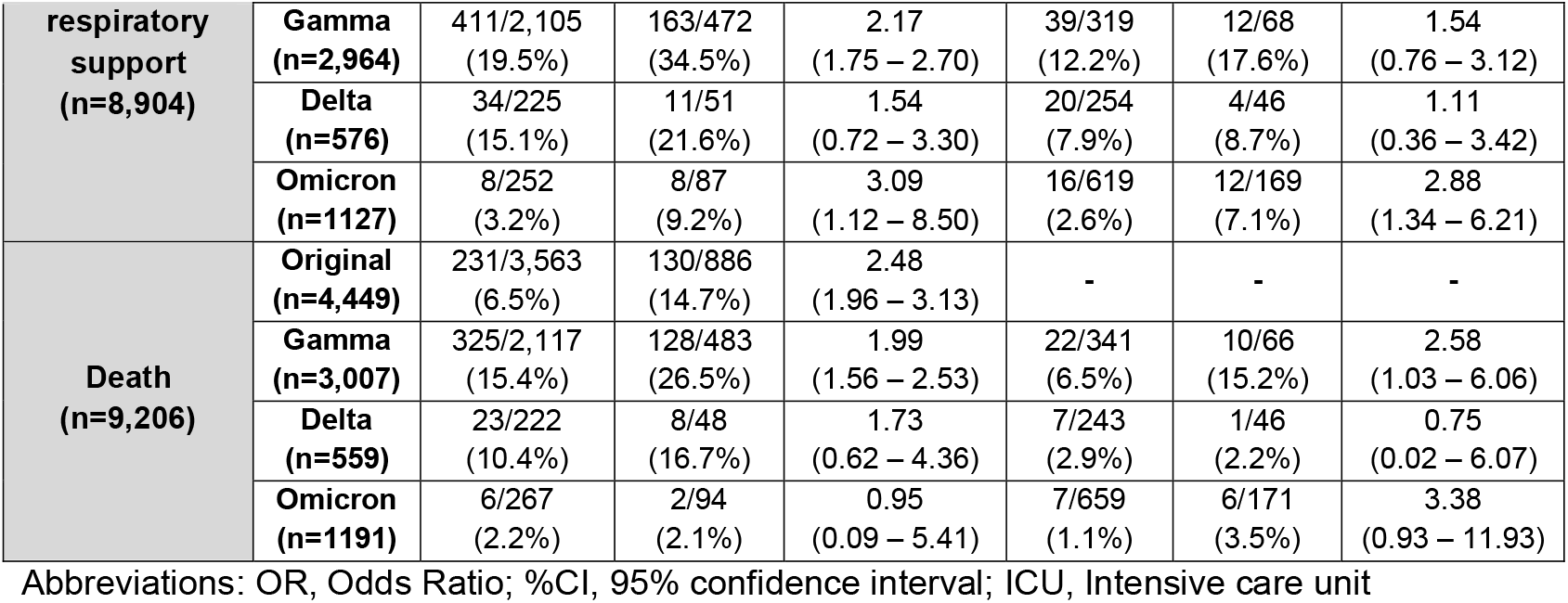
Outcomes of pregnant and postpartum women with COVID-19.

Among vaccinated patients with the Gamma variant, patients in the puerperal group were more likely to die than those in the pregnant group (OR: 2.58; 95% CI: 1.03 – 6.06). Among vaccinated patients with the Delta variant, symptoms, ICU admission, requirement for invasive respiratory support, and death rate were not significantly different between the pregnant and puerperal groups. Patients in the puerperal group with the Omicron variant who were vaccinated had a lower frequency of fever (OR: 0.43. 95% CI: 0.28 – 0.66) and vomiting (OR: 0.35; 95% CI: 0.12 – 0.85) and a higher frequency of dyspnoea (OR: 1.64; 95% CI: 1.07 – 2.50), SpO_2_ < 95% on room air (OR: 1.97; 95% CI: 1.20 – 3.22), ICU admission (OR: 1.91; 95% CI: 1.13 – 3.17), and invasive respiratory support (OR: 2.88; 95% CI: 1.34 – 6.21), compared with those in the pregnant group.

## DISCUSSION

In this study, the clinical findings and outcomes of pregnant and postpartum women with COVID-19 differed. These differences were influenced by the predominant VOCs at the time of diagnosis (original, Gamma, Delta, and Omicron) and patient vaccination status.

### postpartum women

When the COVID-19 pandemic began in 2020, it was believed that pregnant and puerperal women were not at a higher risk of adverse outcomes or death than the non-obstetric population.[3, 4, 24-27] However, updated studies reported higher rates of invasive ventilation, ICU admissions, and death due to COVID-19 in the obstetric population.[6-8, 10, 28, 29] Differences in clinical findings and outcomes between pregnant and postpartum women are unclear, as previous studies regarding these differences included non-hospitalized patients in whom a COVID-19 diagnosis was not confirmed using RT-PCR.[8-11]

Another previous study compared the clinical findings and outcomes of pregnant, puerperal, and non-pregnant nor puerperal women aged 10 to 49 years with COVID-19 from the beginning of the pandemic until 2 January 2021. This previous study used propensity score matching and found that puerperal women were more likely than pregnant women to be admitted to the ICU (OR: 1.97), require invasive respiratory support (OR: 2.71), and die (OR: 2.51).[15]

The three delays model can be used to explain why postpartum women had worse prognoses than pregnant women in this study.[30] During the postpartum period, women may delay seeking medical assistance, as they focus on taking care of their new-born, ignoring their own health care.[31, 32] The risk of thromboembolism increases during the postpartum period as well as in patients with COVID-19.[33-37] As the SIVEP-Gripe does not provide information regarding the date of delivery or the gestational age upon diagnosis, some patients in the puerperal group may have been infected with SARS-CoV-2 during pregnancy that progressed to SARS after delivery. In addition, pregnant women with SARS may terminate their pregnancy as a therapeutic measure and may die during the puerperal period. Caesarean sections account for more than 57% of deliveries in Brazil and are even more frequent among patients with severe COVID-19, which may increase the risk of maternal mortality in this population.[37-40]

Among the unvaccinated patients in this study, those in the puerperal group had worse prognosis than those in the pregnant group for each VOC. In the puerperal group, 35.6%, 48.0%, 51.0%, and 20.5% of unvaccinated patients with the original, Gamma, Delta, and Omicron variants, respectively, were admitted to the ICU; 20.3%, 34.5%, 21.6%, and 9.2%, respectively, required invasive ventilation; and 14.7%, 26.5%, 16.7%, and 2.1%, respectively, died. The mortality rate was significantly lower in unvaccinated pregnant patients, with the exception of patients with the Omicron variant.

Studies including the general population have reported that different VOCs and vaccination statuses affect the clinical symptoms and outcomes of patients with COVID-19.[41-42] A review concluded that the Alpha, Beta, and Gamma variants were associated with higher risks of hospitalization and ICU admission, compared with the original variant and that the Beta variant had the highest risk of ICU admission.[43] A meta-analysis reported higher risks of hospitalization, ICU admission, and mortality for patients infected with the Beta and Delta variants, compared with those infected with the Alpha and Gamma variants. The meta-analysis also concluded that all SARS-CoV-2 VOCs are associated with a higher risk of severe outcomes, compared with the original SARS-CoV-2 variant.[44]

In this study, postpartum patients were less likely to have comorbidities, although the comorbidities varied between the VOC subgroups. Patients in the puerperal group also presented with different clinical findings based on the VOC and their vaccination status. Patients in the puerperal group who were unvaccinated were more likely to have an SpO_2_ < 95% on room air. This was true in all VOC subgroups. Unvaccinated patients in the puerperal group with the original, Gamma, and Omicron were more likely to be admitted to the ICU and to require invasive ventilatory support, and those with the original and Gamma variants were more likely to die.

Among the vaccinated puerperal patients, those in Omicron variant subgroup had a higher risk of having an SpO_2_ < 95% on room air. Vaccinated puerperal patients in the Omicron subgroup also had a higher risk of ICU admission and requiring invasive ventilatory support. Those in the Gamma subgroup had an increased risk of death. These findings suggest that puerperal women have a higher risk of severe outcomes than pregnant women, especially when unvaccinated.

Previous observational studies and surveillance data regarding vaccination during pregnancy are reassuring. Most patients in the previous studies were administered the Pfizer/BioNTech or Moderna vaccines, and no adverse outcomes or side effects of vaccination during pregnancy were reported.[45-48] In Brazil, pregnant women are administered the Sinovac/Butantan and Pfizer/Wyeth vaccines.[49] In hospitalized pregnant and postpartum patients with severe COVID-19, those who received two doses of a COVID-19 vaccine had a 46% reduction in the odds of ICU admission, an 81% reduction in the odds of requiring invasive ventilatory support, and an 80% reduction in the odds of death, compared with those who did not receive any COVID-19 vaccination.[19] Among the general population, several studies have indicated that COVID-19 vaccination protects against severe outcomes caused by SARS-CoV-2 variants, including the Omicron variant.[50-52] The results of the current study support these previous findings and suggest that vaccines effectively reduce adverse outcomes including ICU admission, the requirement for invasive ventilatory support, and death in pregnant and puerperal patients. Therefore, vaccination is an important public health strategy globally, and pregnant and postpartum women should be included in randomized clinical trials.[19, 53].

This study has several strengths, including the use of a large dataset with nationwide coverage and no duplicates that included hospitalized obstetric patients with COVID-19 confirmed via RT-PCR, VOC data, and vaccination status. Similar studies neither reported these data nor divided the obstetric population into pregnant and puerperal groups.

However, this study has limitations, including its retrospective nature based on a secondary database analysis. Although reporting COVID-19-related hospital admissions is compulsory in Brazil, it is not guaranteed that all patients with COVID-19 who were hospitalized were included in the database. Bias due to missing or inaccurate data cannot be ruled out. Asymptomatic or mildly symptomatic patients are not reported in the SIVEP-Gripe; therefore, an analysis of the non-hospitalized population was not possible. In addition, the database used in this study does not include information regarding other obstetric variables such as gestational age, delivery mode, date of birth, comorbidities, week of pregnancy at diagnosis, and perinatal data. As it is an anonymous database, the data cannot be linked with the public database of birth registers and mode of delivery.

As postpartum women have higher risks of developing severe forms of COVID-19, especially when unvaccinated, health care strategies to implement vaccination and manage postpartum patients with COVID-19 are necessary. The risk of severe COVID-19 in puerperal patients should not be underestimated, and health care workers should closely monitor all postpartum women infected with SARS-CoV-2, especially those who are not vaccinated. The vaccination of pregnant and postpartum patients should become a public health priority.

## Data Availability

The data that support the findings of this study are available in GitHub repository at https://github.com/observatorioobstetrico/COVID-19-impact-of-original-Gamma-Delta-and-Omicron-variants-of-SARS-CoV-2-in-vaccinated-and-unv. These data were derived from the following resources available in the public domain: https://opendatasus.saude.gov.br/dataset/srag-2020 and https://opendatasus.saude.gov.br/dataset/srag-2021-e-2022 obtained on 25 July 2022.

## Acknowledgements

None

## Competing interests

The authors have declared that no competing interests exist.

### Funding

This work was supported, in whole or in part, by the Bill & Melinda Gates Foundation [INV-027961]. Under the grant conditions of the Foundation, a Creative Commons Attribution 4.0 Generic License has already been assigned to the Author Accepted Manuscript version that might arise from this submission. This work is also funded by CNPq (Award Number: 445881/2020-8) and FAPES (Award Number: 44437.699.40413.22122020). The funders had no role in study design, data collection and analysis, decision to publish, or preparation of the manuscript.

